# Ten years malaria trend analysis in Mangili Public Health Centre, East Sumba District of East Nusa Tenggara Province, Indonesia: a retrospective study

**DOI:** 10.1101/2023.11.18.23298721

**Authors:** Maria Lobo, Robertus Dole Guntur, Damai Kusumaningrum, Yulianti Paula Bria

**Author notes:** **Address for Correspondence:** Ir. Maria Lobo, M.Maths.Sc., PhD, Affiliation: Mathematics Study Program Faculty of Science and Engineering, Nusa Cendana University Jln. Adisucipto Penfui Kupang NTT, Indonesia Post Code 85001.

## Abstract

**BACKGROUND:** Malaria still remains a major public health problem globally including in Indonesia. Currently, most of malaria cases are in the eastern part of the country. However, there is no information yet regarding the trends of malaria prevalence at rural health institution particularly at Mangili Public Health Centre.

**AIM:** This study aims to explore the trend of malaria prevalence over a ten-year period.

**METHODS:** A retrospective study was carried out in the Centre. The laboratory record books from January 2013 to December 2022 were carefully reviewed to identify malaria cases based on the sex, age, type of plasmodium, year and month when the patients diagnosed malaria microscopically. The trend analysis was applied to identify the trend of malaria over the period under review.

**RESULTS:** Over the last ten years, 19,938 blood films were requested for malaria diagnosis at the health centre. From this number, 3.679 (18.5%, with 95% confidence interval (CI): 17.2 – 19.7) were microscopically confirmed malaria cases. From the total cases, prevalence of plasmodium falciparum, vivax and mix infection was 85.5% with 95% CI: 84.3 – 86.8; 6.39% with 95% CI: 3.26 – 9.51; and 7.58% with 95% CI: 4.48 – 10.7 respectively. The number of malaria cases reached a peak in January, whilst it was the lowest in October. Overall trend on the prevalence of malaria decreased significantly, however the prevalence of plasmodium falciparum increased considerably during COVI-19 pandemic. The prevalence of plasmodium falciparum, vivax and mixed infection was higher in male, age group ≥ 15, and student than their counterpart.

**CONCLUSION:** Malaria remains high in this area with plasmodium falciparum is the dominant species followed by mixed infection. Scaling up malaria control and prevention activities are very crucial to significantly reduce the burden of malaria and to boost malaria elimination in this region.

## Introduction

Malaria is a significant global health issue distributing across 84 malaria endemic nations (1) and leading for causes of death in many developing countries (2). The current data of World Health Organization (WHO) indicates that the number of malaria cases in 2021 was about 247 million. The majority of the cases (95%) was in African countries, whilst other cases were in other countries including 2 % from nations in the region of South-East Asia (SEA). The region’s action plan indicates that by 2030, the entire region will be declared a malaria-free zone (3). Of the 11 nations in this area, the WHO has provided a certificate for malaria-free status to both Maldives and Sri Lanka. One indicator compulsory to accomplish the certificate is that there should have been no local malaria cases over the preceding three consecutive years (4). The highest burden of malaria in this region in 2021 was in India and Indonesia, accounting to 79.2% and 16.6% of the region’s total cases, respectively (1).

Indonesia which has 38 provinces administratively has a national commitment to achieve a malaria elimination area by 2030 (5). All districts in the country have been classified as malaria free zone, low, moderate, and high malaria endemic settings (MES) based on their annual parasite incidence (API). API was defined as the number of malaria cases confirmed microscopically per thousand people (5). The current API indicates that in 2021, the national API was 1.1 per 1,000 population and this figure differs throughout the country (6). Most of districts in the western part of the country has API value less than 0.05 and have been categorized as malaria free zone, whilst in the Eastern part of the country, most of districts have API value more than 0.1 including the top three of highest value of API was in Papua, West Papua and East Nusa Tenggara (ENT) Province with the value 80.5, 7.56, and 1.69 per thousand population respectively (6).

ENT Province is an archipelago province consisting of five main islands including Flores, Timor, Lembata, Alor and Sumba [11]. The total number of malaria cases in 2022 was 15,830 cases with most of the cases (84%) was in the Sumba Island (7). The island consists of four districts which three of them, Southwest Sumba, East Sumba, West Sumba districts are categorised as high malaria endemic settings (MES). Almost 50% of malaria cases in Sumba Island was contributed from the Eastern part of the island (7). The cases were scattered in all sub-district of East Sumba district allowing 100% of 248,776 total population in the region could have a risk of contracting the diseases (8). The rate of malaria risk and transmission vary between season (9) with the highest in December and the lowest in April (10). Moreover, the incidence and prevalence of malaria differs depending on environmental and socio-demographic factors including gender and age group (11). Therefore, monitoring the burden of malaria and the trend in endemic area are essential to examine the measures and the impact of any intervention. However, such useful data remain limited in several endemic areas of Eastern part of Sumba.

Risk of malaria was high in East Sumba district, in which all sub-districts are prone to malaria transmission. For instance, analysis of one year data of malaria patients in 2015 admitted in Public Hospital of Umbu Rara Meha in the capital city of East Sumba district revealed that prevalence of plasmodium falciparum was higher than plasmodium vivax and more than half of the cases was contributed by Waingapu city (12). Another cross-sectional study of 262 patients admitted at Lindimara Hospital of East Sumba from July to December 2014 indicated that the prevalence of malaria in female was higher than in male with most of the patient suffered with single infection of plasmodium falciparum (13). Then, a retrospective study to analyse two year trend of malaria in East Sumba district revealed that the prevalence of malaria in 2018 and 2019 was higher in male than female (14). Strikingly, a total of 5,537 malaria cases were recorded in East Sumba District from Jan 2021 to December 2022 (7). In spite of the general public health significance and the widespread incidence of malaria in all sub-districts of East Sumba, the general trend of malaria prevalence in the last ten year has not been investigated. The evaluation of morbidity pattern of malaria in endemic areas would help policy maker to understand the dynamic of diseases transmission and to design the effective intervention tailored with local condition to boost malaria elimination in the district. Furthermore, there is no information yet regarding the trends of malaria prevalence at the health centres and particularly at Mangili Public Health Centre in the last ten year. Therefore, the aim of this study was to evaluate the trend of malaria and transmission pattern regard sociodemographic factors and season in the last ten year at Mangili Public Health Centre of East Sumba. Therefore, the present study addressed the above gaps with the hope to provide a significant contribution to boost the achievement of malaria elimination in East Nusa Tenggara Province Indonesia.

## Materials and Methods

### Study settings

A retrospective study was performed from August to October 2023 to determine a ten-year trend analysis of malaria prevalence in the working areas of Mangili Public Health Centre by assessing malaria blood film of malaria patients admitted in the centre from January 2013 to December 2022. This health centre is situated on the Pahunga Lodu Sub-district covering eight villages. The sub-district has a total area of 349.8 kilo metre. The location of the sub-district is about 119 km in the Eastern part of Waingapu city, having an elevation of 216 meters above sea level. Based on the 2020 census conducted by Central Bureau Statistics of East Sumba District, the sub-district has a total population of 13,066 of whom 51% are males and 49% females (15)

### Data collection procedure

Medical record of malaria patients registered at the Mangili Public Health Centre from January 2013 to December 2022 was investigated in the coordination between the head of malaria section of Health Department of East Sumba District and the Head of Mangili Public Health Centre. In collaboration with the malaria program manager of the centre, individual data such as total clinically treated, confirmed case in month and year, types of malaria species and socio-demographic data (age, sex) was composed. Any data such as the socio demographic, malaria diagnosis result which is not properly recorded was excluded.

### Statistical analysis

All data was verified for its completeness. The retrospective malaria data was recapitulated using figures and tables. Specifically, the total malaria case in the past ten years (from January 2013 to December 2022), malaria case by age and sex was summarized using table. Socio-demographic data of participants was described by descriptive statistics. The proportion of malaria infection in each year and its 95% confidence interval (CI) was computed from 2013 to 2022. Furthermore, trend analysis was used to compare plasmodium types composition in each year and to assess seasonal transmission of malaria during the period under review. Data was examined using SPSS.

### Ethics approval and consent to participate

This study was approved by Health Research Ethics Committee of Nusa Cendana University (reference number 51/UN15.16/KEPK/2023). Permission letter was further obtained from the governor of ENTP, regent of East Sumba District, and Health department of East Sumba District. The head of Mangili Public Health Centre had been provided with sufficient information in relation with the purposes, risk, and advantage of the study before the research team start to collect data.

## Results

### Retrospective malaria data

A total of 19,938 blood films were prepared and scrutinized from malaria suspected patients at the Mangili Health Centre from January 2013 to December 2022. Of the total examined blood films, 18.5% (3,679) with 95% confidence interval (CI): 17.2 – 19.7 were confirmed malaria cases microscopically. The highest number of malaria-suspected patients was in 2014 with 3227 patients, whilst the lowest was in 2016 with 1,067 patients. From the medical record, it was found that the highest number of confirmed malaria cases was observed in 2014, accounting for 1397 positive cases. Overall, there were a significant reduction in the trend of malaria prevalence over the ten-year period at Mangili Health Centre, from 31.7% with 95% CI: 28.4 - 35.0 at the beginning of the period (2013) to 6.08% with 95% CI: 2.48 - 9.69 at the end of period (2022). The highest malaria prevalence was recorded in 2014 at 43.3% with 95% CI: 40.7 – 45.9, meanwhile it was the lowest in 2022 at 6.08% with 95% CI: 2.48 – 9.69.

Regarding to the plasmodium species of the patients, it was found that the majority of the cases in Mangili Public Health Centre was due to single infection of *plasmodium falciparum*, accounting for 85.5% with 95% CI: 84.3 – 86.8, followed by mixed infection (*plasmodium falciparum* and plasmodium vivax) and *plasmodium vivax* single infection accounted for 7.58 % with 95% CI: 4.48 – 10.7 and 6.39% with 95% CI:3.26 – 9.51 respectively. The trend of *plasmodium falciparum* prevalence decreased slightly from 98.2% with 95% CI :97.2 – 99.1 at the initial point in 2013 to 80.5% with 95% CI: 73.8 – 87.1 at the final point of the review (2022). The highest prevalence of single infection of *plasmodium falciparum* was observed in 2013, whilst it was the lowest in 2020 at 39.9% with 95% CI: 27.8 – 52.0. Furthermore, the highest prevalence of single infection of plasmodium vivax was noticed in 2017 at 9.78% with 95% CI: 0.57 – 19.0, whilst for mixed infection due to plasmodium falciparum and plasmodium vivax, the highest prevalence was in 2020 at 47.5% with 95% CI: 36.2 – 58.8. It is interesting to note that during the COVID-19 period (from 2020 to 2022) the prevalence of single infection due to plasmodium falciparum shows an increasing trend from 39.9% with 95% CI: 27.8 – 52.0 in 2020 to 80.5% with 95% CI : 73.8 – 87.1 in 2022, meanwhile the prevalence of mixed infection due to mixed infection of plasmodium falciparum and vivax depicted the downward trend from 47.5% with 95% CI: 36.2 – 58.8 in 2020 to 13.0% with 95% CI : 0.00 – 27.1 in 2022 as indicated in Table 1.

**Table 1.** Profile of ten-year malaria cases report from Mangili Health Centres of East Sumba District of East Nusa Tenggara from 2013 to 2022.

| Year | Total of blood films examined | Total number of cases |  | *Plasmodium falciparum* |  | *Plasmodium vivax* |  | *Mix (Pf + Pv)* |  |
| --- | --- | --- | --- | --- | --- | --- | --- | --- | --- |
|  |  | n | % (95% CI) | n | % (95% CI) | n | % (95% CI) | N | % (95% CI) |
| 2013 | 2428 | 770 | 31.7 (28.4, 35.0) | 756 | 98.2 (97.2, 99.1) | 13 | 1.69 (0.00, 8.69) | 1 | 0.13 (0.00, 7.19) |
| 2014 | 3227 | 1397 | 43.3 (40.7, 45.9) | 1310 | 93.8 (92.5, 95.1) | 75 | 5.37 (0.27, 10.5) | 0 | 0.00 (0.00, 0.00) |
| 2015 | 1263 | 195 | 15.4 (10.4, 20.5) | 171 | 87.7 (82.8, 92.6) | 24 | 12.3 (0.00, 25.5) | 0 | 0.00 (0.00, 0.00) |
| 2016 | 1067 | 168 | 15.7 (10.2, 21.3) | 149 | 88.7 (83.6, 93.8) | 11 | 6.55 (0.00, 21.2) | 8 | 4.76 (0.00, 19.5) |
| 2017 | 1701 | 409 | 24.0 (19.9, 28.2) | 273 | 66.7 (61.2, 72.3) | 40 | 9.78 (0.57, 19.0) | 94 | 23.0 (14.5, 31.5) |
| 2018 | 1859 | 133 | 7.15 (2.77, 11.5) | 107 | 80.5 (72.9, 88.0) | 10 | 7.52 (0.00, 23.9) | 14 | 10.5 (0.00, 26.6) |
| 2019 | 2231 | 192 | 8.61 (4.64, 12.6) | 121 | 63.0 (54.4, 71.6) | 23 | 12.0 (0.00, 25.3) | 48 | 25.0 (12.8, 37.3) |
| 2020 | 2097 | 158 | 7.53 (3.42, 11.7) | 63 | 39.9 (27.8, 52.0) | 20 | 12.7 (0.00, 27.2) | 75 | 47.5 (36.2, 58.8) |
| 2021 | 1287 | 88 | 6.84 (1.56, 12.1) | 61 | 69.3 (57.7, 80.9) | 8 | 9.09 (0.00, 29.0) | 17 | 19.3 (0.55, 38.1) |
| 2022 | 2778 | 169 | 6.08 (2.48, 9.69) | 136 | 80.5 (73.8, 87.1) | 11 | 6.51 (0.00, 21.1) | 22 | 13.0 (0.00, 27.1) |
| Total | 19938 | 3679 | 18.5 (17.2, 19.7) | 3147 | 85.5 (84.3, 86.8) | 235 | 6.39 (3.26, 9.51) | 279 | 7.58 (4.48, 10.7) |

Abbreviation: *Pf* = *Plasmodium falciparum*, *Pv* = *Plasmodium vivax*

### Seasonal Variation of Plasmodium Types

The trend of the number of malaria cases based on season and month in the last ten year was presented in Figure 1. Overall, the number of malaria cases has been reported in all month and season. The outcome of this study indicated that the highest number of the cases was in January, accounting for 583 cases, whilst it was the lowest in October with 147 cases. The transmission of malaria shows an increasing trend from the beginning of rainy season in October until reach a peak in January, then it decreased gradually until the end of rain season in April. During the transition period from rainy season to dry season, the number of malaria cases rose gradually from 323 cases in April to 397 cases in May. After that the number of malaria cases decreased until the end of dry season. In relation to the plasmodium types, this study depicted that the number of single infection due to plasmodium falciparum showed a decreased trend during the dry season, whilst it was increased significantly during the rainy season. The trend of malaria cases due to single infection of plasmodium vivax and mixed infection (due to vivax and falciparum) do not show fluctuated throughout the year, however the maximum point was reached in different time. The highest number of mixed infection (plasmodium falciparum and plasmodium vivax) was in January, whilst it was in March for plasmodium vivax as shown in Figure 1.

**Figure 1:**
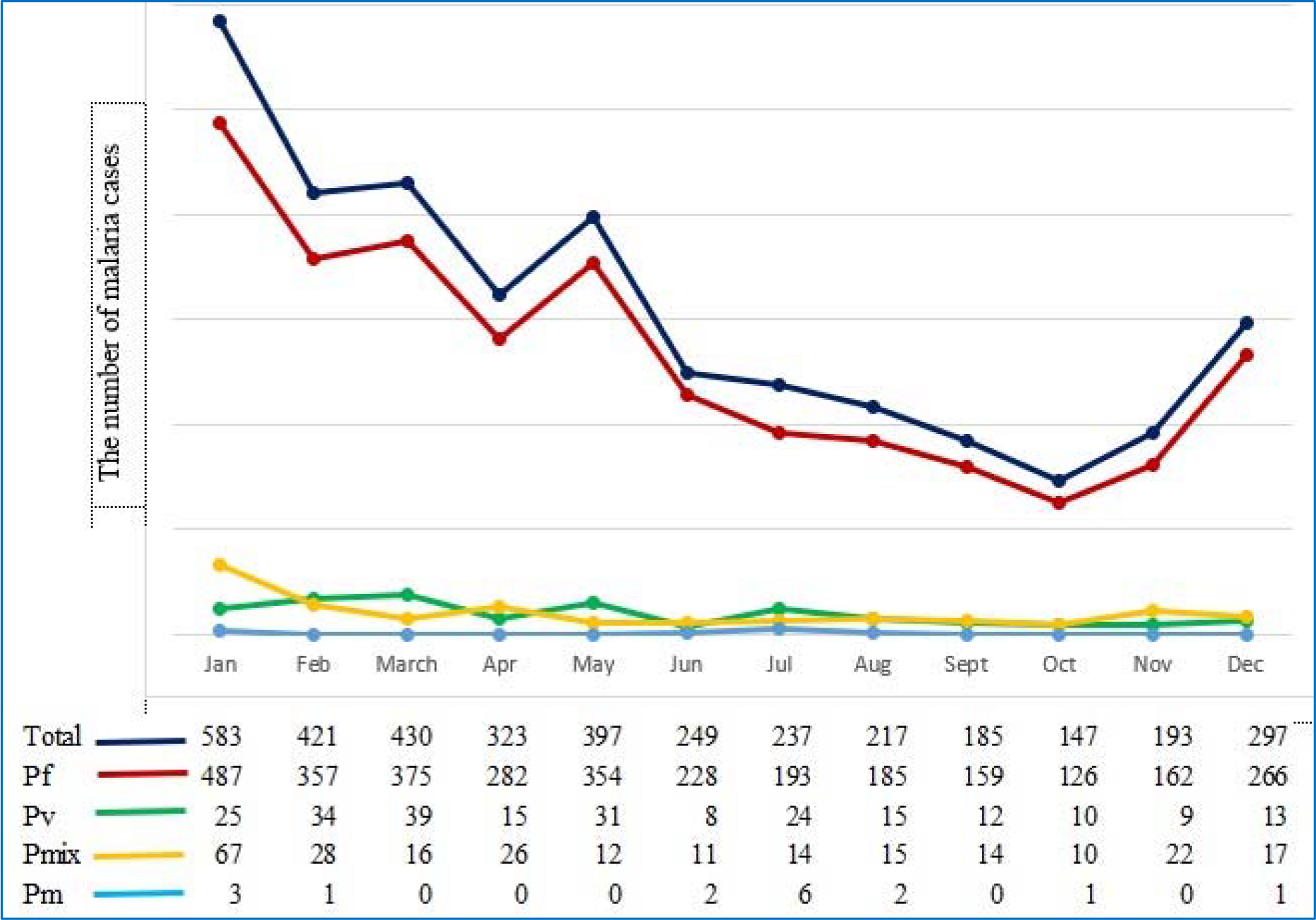
Monthly trends of malaria for a ten-year period at Mangili Health Centre from 2013 to 2022

### Trend of malaria cases by age group

Based on the record review over the ten years from 2013 to 2022 in the location of the study, it was found that malaria was reported in all age group. The highest burden of malaria in each year over the period under review was in adult group which has age more than 15 years old. The highest number of malaria cases in the group of children less than five year, 5 to 9 year, 10 to 14 year, and adult group was observed in 2014, contributing to 207 cases, 379 cases, 279 cases 532 cases respectively. After the period of 2014, the number of malaria cases decreased for all age group until the end the period as indicated in Figure 2.

**Figure 2:**
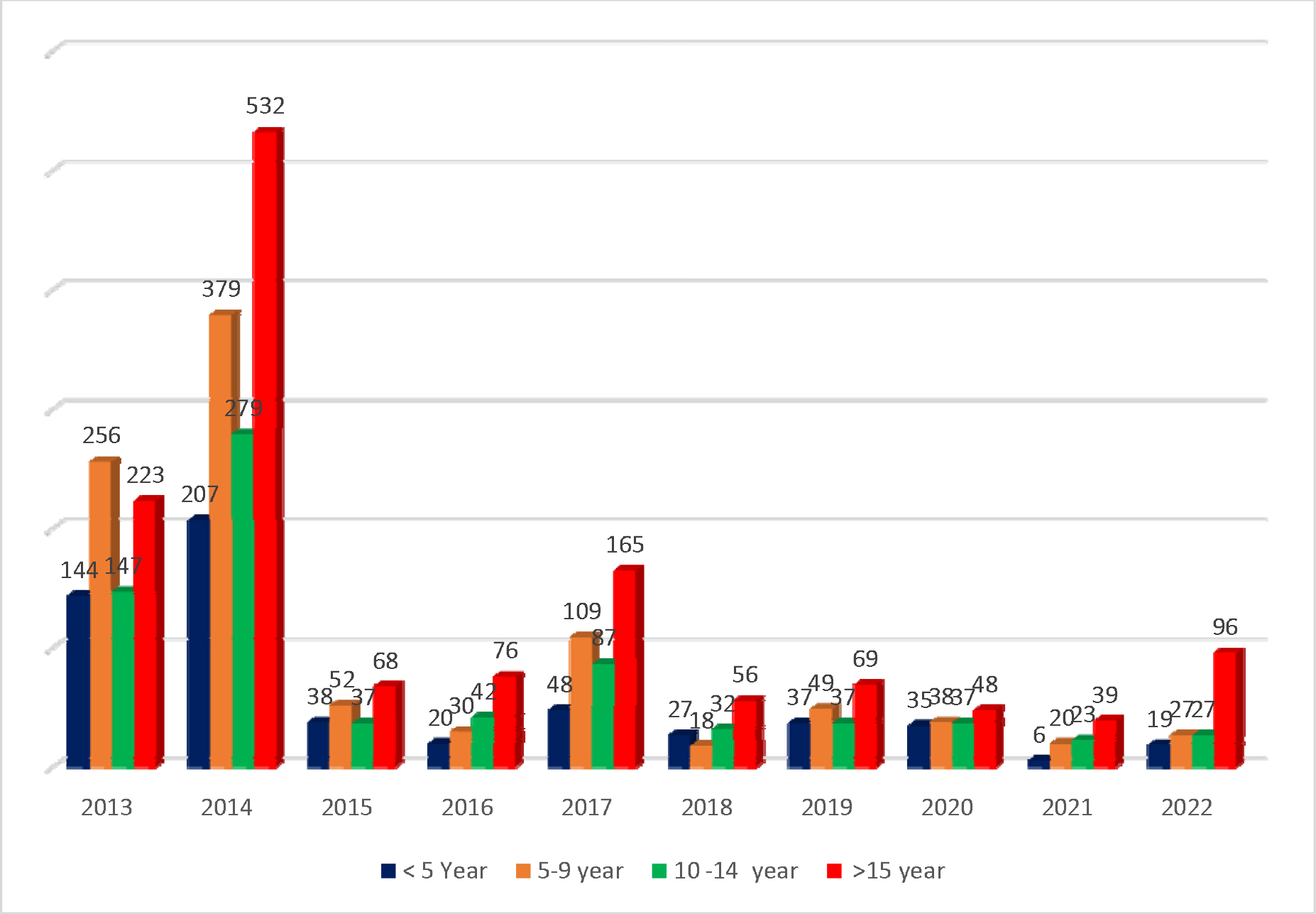
Trends of the number malaria patients based on the age group for a ten-year period at Mangili Health Centre from 2013 to 2022

### Trend of malaria case in gender

During the period from 2013 to 2022, the number of malaria infection in male was higher than in female except in 2018. In that year, the number of cases between male and female almost comparable. The number of malaria cases in male in 2013 was 401 cases. This figure was almost double in 2014, accounting for 744 cases. After that the cases in the male group decreased significantly to 85 cases in 2016. This figure rose sharply to 210 cases in 2017, after that the number of cases in male group decreased until the end of period under review. The number of malaria cases in female group also provided a downward trend with the highest cases was in 2014, accounting for 653 cases and the lowest number was in 2021 with 39 cases as shown in Figure 3.

**Figure 3:**
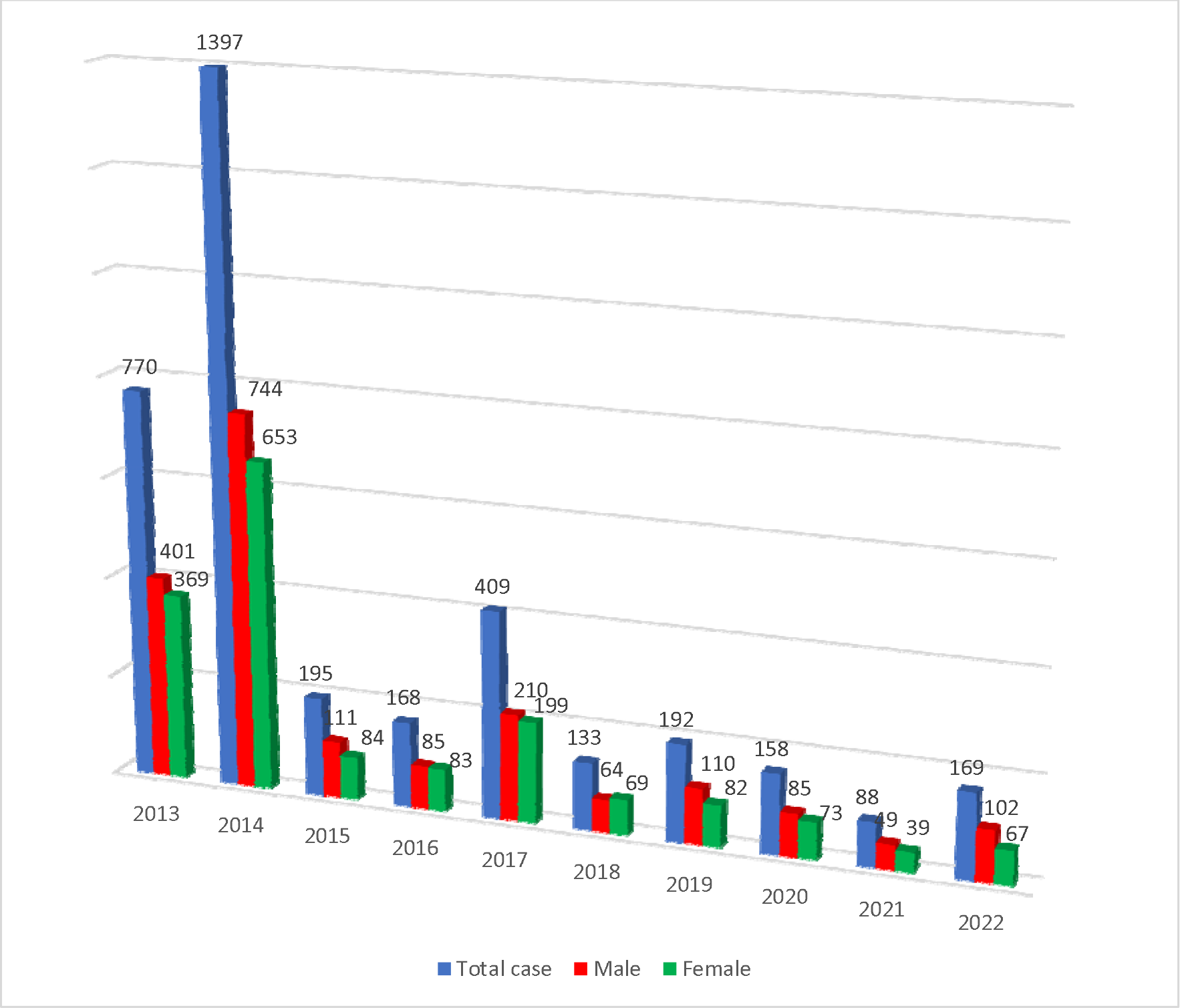
Distribution of malaria cases by gender in Mangili Health Centre of East Sumba District of East Nusa Tenggara for a ten-year period from 2013 to 2022

### Trend of malaria cases by socio-demographic

The distribution of malaria patients and plasmodium species based on socio-demographic and other variables in Mangili Public Health Centre for the four-year period from 2019 to 2022 was displayed in Table 2. Overall, the number of malaria cases reported in the centre during the period under review was 605 cases. The prevalence of malaria was higher in male than in female (56.9% vs 43.1%). Regarding to age group, the prevalence of malaria in adult group (people with age more than 15 years old) was the highest (41.5%) of other age groups. Almost half of the malaria patients in this health centre was working as student (47.8%). There were ten patients who were pregnant suffered from malaria during the period under review. Almost all malaria cases (95.5%) in Mangili Health Centre were found during the patient attending the centre. Other cases were identified when malaria section of the centre conducted some programs including visiting home, contact survey and mass blood survey, accounting for 2.81% (17 cases), 1.32% (8 cases), and 0.33 (2 cases) respectively.

**Table 2.** Distribution of plasmodium species by socio-demographic in Mangili Health Centre for a four-year period from 2019 to 2022.

|  | Plasmodium types |  |  | Total |
| --- | --- | --- | --- | --- |
|  | Pf | Pv | PMix |  |
| **Total** | **381** | **62** | **162** | **605** |
| **Gender** |  |  |  |  |
| Male | 219 (57.5) | 31 (50.0) | 94 (58.0) | 344 (56.9) |
| Female | 162 (42.5) | 31 (50.0) | 68 (42.0) | 261 (43.1) |
| **Age group** |  |  |  |  |
| < 5 | 54 (14.2) | 11 (17.7) | 32 (19.8) | 97 (16.0) |
| 5 - 9 | 83 (21.8) | 16 (25.8) | 35 (21.6) | 134 (22.1) |
| 10 - 14 | 74 (19.4) | 10 (16.1) | 39 (24.1) | 123 (20.3) |
| >=15 | 170 (44.6) | 25 (40.3) | 56 (34.6) | 251 (41.5) |
| **Main Job** |  |  |  |  |
| Housewife | 49 (12.9) | 9 (14.5) | 13 (8.02) | 71 (11.74) |
| office staff | 6 (1.57) | 1 (1.61) | 3 (1.85) | 10 (1.65) |
| Student | 180 (47.2) | 29 (46.8) | 80 (49.4) | 289 (47.8) |
| Farmer | 75 (19.7) | 9 (14.5) | 25 (15.4) | 109 (18.0) |
| No working | 71 (18.6) | 14 (22.6) | 41 (25.3) | 126 (20.8) |
| **Pregnant status** |  |  |  |  |
| Yes | 3 (0.79) | 3 (4.84) | 4 (2.47) | 10 (1.65) |
| No working | 378 (99.2) | 59 (95.2) | 158 (97.5) | 595 (98.3) |
| **Cases finding** |  |  |  |  |
| Health facilities | 359 (94.2) | 57 (91.9) | 162 (100) | 578 (95.5) |
| Visiting home | 14 (3.67) | 3 (4.84) | 0 (0) | 17 (2.81) |
| Contact survey | 8 (2.1) | 0 (0) | 0 (0) | 8 (1.32) |
| Mass blood survey | 0 (0) | 2 (3.23) | 0 (0) | 2 (0.33) |

Turning to the plasmodium species, this study found that the prevalence of malaria due single infection of plasmodium falciparum and plasmodium vivax was 63% (381/605) and 10.2% (62/605) respectively. Whilst the prevalence of malaria due to mixed infection of plasmodium falciparum and plasmodium vivax was 26.8% (162/605). From the total number of single infections of plasmodium falciparum and mixed infection, its prevalence was higher in male than in female group (57.5% vs 425%, 58% vs 42% respectively). Whilst the prevalence of single infection due to plasmodium vivax was the same between male and female group.

The distribution of plasmodium types in Mangili Health Centre indicated that the prevalence of each plasmodium types varies significantly amongst age group. The highest prevalence of single infection due to plasmodium falciparum, plasmodium vivax and mixed infection was in the age group of adults, accounting for 44.6%, 40.3%, and 34.6% respectively. This study showed that the prevalence of mixed infection was in line with the increase of the age of the patients with the lowest was in children less than five and the highest was in the adult groups. In relation to the main job of the patients, the prevalence of each type of infection shows a difference amongst diverse occupation. The highest prevalence of single infection due to plasmodium falciparum, vivax, and mixed infection was in the group of students, accounting for 47.2%, 46.8%, and 49.4% respectively as shown in Table 2.

## Discussion

To the best our knowledge this is the first study to investigate the trend of plasmodium falciparum and plasmodium vivax using large database from the local health centre in rural East Nusa Tenggara Province Indonesia for the last ten year. This study found that in Mangili Public Health Centre the prevalence of malaria was high in the last ten year, however, the overall trend of malaria prevalence decreased significantly during the period under review. Furthermore, the prevalence of malaria falciparum was higher than malaria vivax and the number of malaria cases reached a peak in January and the lowest was in October. During the COVID-19 pandemic, the prevalence of single infection due to plasmodium falciparum rose significantly.

This study showed that the prevalence of plasmodium falciparum was higher than plasmodium vivax. This consistent with the national report of Indonesia government (5) and study in many parts of Indonesia (14,16–18) and other countries including Ethiopia (19), Northwest Ethiopia (20) Saudi Arabia (21), but this contrast with study in Mauritania (22) indicating the prevalence of malaria vivax was higher than falciparum. In this study, the prevalence of plasmodium falciparum shows a decreased trend from year to year, however the number of the cases was still high until the end of period under review. Stakeholders should provide much attention to tackle plasmodium falciparum, due to its clinical complications and drug-resistant threat in this archipelago (23). Interestingly, the prevalence of single infection due plasmodium vivax and the prevalence of mixed infection due to plasmodium falciparum dan plasmodium vivax was high in the last ten year. The high number of malaria vivax found in this study is a signal for the local authority that more attention should be paid for this situation as treatment of plasmodium vivax need higher awareness of local community to take the medication for fourteen days to achieve to optimal result in curing the malaria vivax (24–26).

This study shows that the number of malaria cases shows an increasing trend during COVID-19 pandemic. This finding was consistent the global trend of malaria due to this pandemic (27) and other findings in Saudi Arabia (21) and Zimbawe (28). This could be understood as during this pandemic time, the access to health service was limited which might impact on health-seeking behaviour of local community. Fear of infection to the COVID-19 leads to people do not seek treatment for their malaria on the health facilities, as a result malaria patient was not diagnosed and do not receive the proper treatment. This could lead to local transmission of malaria continue in the community contributing to increasing the number of malaria cases. However, it is worth to note that the increasing number of malaria cases in this region was not entirely due to the disruption during COVID-19. For instance, environmental factors such as temperature and rainfall might be contributed to the higher occurrence of malaria in the nation. Previous study has showed the important role of environmental factors play in malaria transmission in Indonesia (29,30). To better understand the factors that trigger high number of malaria cases during the pandemic, further investigation of the impact of the COVID-19 to this region is needed.

This study further shown that in the study area, the number of malaria cases peaked in wet season and the lowest malaria cases was in the beginning of transition period from rainy to dry season in period of April to Mei. This finding corroborates with other result in other settings (31–33). This could be understood as during the wet season from October to April, the habitat of mosquito might be extended (34) and there were many puddles which have the highest larva density amongst other breading site of mosquitos (35). This condition could provide an appropriate environment for Anopheles mosquito to bread and grow well. Therefore, the transmission of malaria run well to achieve the highest number of the cases during this wet period. Whilst during dry season from April to September, the minor transmission period might occur as a result of low rainfall. Although malaria transmission is usually associated with rainy seasons, in this study malaria cases were also significant in dry season, indicating that climatic and environmental factors other than rainfall could also determine the occurrence of malaria in this region.

This study further shown that the prevalence of malaria infection in male group was higher than in female. This finding was consistent with other studies in Sumba Island (10,14) and other parts of Indonesia (36) and other countries including Ethiopia (19) Malaysia (37) indicating that the likelihood to have malaria in the group of male was higher than their counterpart. This could be attributed by life style and main occupation. Most of community around the study area was working in agricultural sector (38) and with the fact that males were mainly engaged in agricultural activities and other large projects. This enables male group was exposure more with malaria vector allowing them have the high risk to have malaria.

Furthermore, our study shows that prevalence of malaria in student was the highest of other occupation. This finding corroborates result in other settings indicated that the prevalence of malaria amongst school age group was higher than their counterpart (39). The high number of malaria cases on the student group might be attributed by the low level of malaria awareness of rural adults in this region. The previous study shown that most of rural adult in East Nusa Tenggara Province had poor malaria awareness indicating that rural adults could not have enough knowledge on how to prevent and treat malaria (24,25,40,41). As a result, rural adults could not act as a row model for their children on how to practice some measures to prevent malaria and to find appropriate treatment when their family members obtained the symptoms of the disease. This situation was worsen by the fact that the prevalence of out of school children in this province was high (42,43). Meanwhile, literature indicated that student in primary school in rural settings could become the best media to spread malaria information on basic malaria knowledge, prevention measures and appropriate treatment seeking behaviour (44). Therefore, the local authority should provide special intervention to enhance malaria awareness of the student in this area to boost malaria elimination in this province.

The strength of the study lies on the fact that malaria data was collected by trained laboratory technician having experience in their field and working as full-time position at the Mangili Health Centre. Furthermore, some malaria data was extracted directly from the Electronic Malaria Surveillance Information System managed by Health Ministry of Indonesia government. However, this study has some limitations including the identification of malaria cases based on the microscopic examination which its sensitivity rate was lower than polymerase chain reaction to identify plasmodium species in the health centre. Then the recorded data in malaria register book was incomplete, therefore the demographic profile of the plasmodium species was merely identified for four years. However, this study provided the baseline data on the malaria prevalence research to be carried out in the future in the Sumba Island of East Nusa Tenggara Province Indonesia.

## Conclusion

In this present study the overall trends of the malaria prevalence decreased significantly in the past ten years from 2013 to 2022, however its prevalence was still high, therefore malaria is still among the major public health problems in Mangili Public Health Centre. Single infection due to plasmodium falciparum is the dominant species in the study area followed by mixed infection due to plasmodium falciparum and vivax. Scaling up malaria control and prevention activities, enhancing malaria campaign for improving malaria awareness of students are very crucial to significantly reduce the burden of malaria and to boost achieving of malaria elimination in the study area.

## Data Availability

All data produced in the present work are contained in the manuscript

## Acknowledgment

We thank all parties for your contribution to this project. We would also like to express our gratitude to the governor of the ENTP, the head of East Sumba, the head of Health Department of East Sumba District, the head of Mangili Public Health Centre and the staff in Malaria section of Mangili Health Centre. Special thank goes to Mrs. Martha Day Mbana as the Manager of Malaria Program in the centre.

